# SARS-CoV-2 antibodies rapid tests: a valuable epidemiological tool in challenging settings

**DOI:** 10.1101/2021.05.08.21256893

**Authors:** Francesca Saluzzo, Paola Mantegani, Valeria Poletti De Chaurand, Virginia Quaresima, Federica Cugnata, Clelia Di Serio, Aurélien Macé, Margaretha De Vos, Jilian A. Sacks, Daniela Maria Cirillo

## Abstract

**Background:** During the last year, mass screening campaigns have been carried out to identify immunological response to SARS-CoV-2 and establish a possible seroprevalence. The obtained results gained new importance with the beginning of SARS-CoV-2 vaccination campaign, as the lack of doses has persuaded several countries to introduce different policies for individuals who had a history of COVID 19.

LFAs may represent an affordable tool to support population screening in LMICs, where diagnostic tests are lacking, and epidemiology is still widely unknown. However, LFAs have demonstrated a wide range of performance and the question of which one could be more valuable in these settings still remains.

**Methods:** We evaluated the performance of 11 LFAs in detecting SARS-CoV-2 infection, analysing samples collected from 350 subjects. In addition, samples from 57 health care workers collected at 21-24 days after the first dose of Pfizer-BioNTech vaccine were also evaluated.

**Findings:** LFAs demonstrated a wide range of specificity (92.31% to 100%) and sensitivity (50 to 100%). The analysis of serum samples post vaccination was used to describe the most suitable tests to detect IgG response against S protein RBD. History of TB therapy was identified as a potential factor affecting the specificity of LFAs.

**Conclusions:** This analysis identified which LFAs represent a valuable tool not only for the detection of prior SARS-CoV-2 infection, but also to detect IgG elicited in response to vaccination. These results demonstrated that different LFAs may have different applications and the possible risks of their use in high TB burden settings.

## Introduction

An accurate knowledge of the local epidemiology of SARS CoV-2 has proven itself crucial to menage the different phases of the COVID 19 pandemic during last year and the possibility to rely on consistent epidemiological data could prove itself helpful to take several public health decisions, also related to the COVID 19 vaccination campaign and WHO COVAX programme. ^1,2^ In the current scenario, in which the lack of vaccines’ doses has persuaded several countries to introduce different policies for individuals who had a history of SARS CoV-2 infection (decision that has not been fully endorsed by WHO) ^3^, the access to reliable data about the serological status of individuals could gain new importance ^4^. However, whereas serological mass screening in high income countries could be feasible using automatic, high-throughput technologies^5,6^, this may not be a practical option in several challenging diagnostic settings, where the prevalence of SARS CoV-2 infection is still widely unknown.^7^ In these countries, lateral flow assays (LFAs) for the identification of SARS CoV-2 antibodies may represent an affordable and practical tool to perform epidemiological evaluations, and a few serological surveys have started to be conducted employing LFAs associated or not with Enzyme Linked Immunosorbent Assays (ELISAs).^8,9^

Even if the numerous SARS-CoV-2 antibody-detection LFAs available on the international market have demonstrated a varying performance^10-13^, generally this test type is considered homogenously, without taking in account their own different characteristics. This fact contributed to generate a common feeling of distrust toward them in the scientific community^14^ and a consensus on which LFAs could be employed as effective epidemiological tools has not been reached. To date, the World Health Organization (WHO) recommends their use only in research for a possible epidemiological employment,^15^ and no LFA has received WHO Emergency Use Listing.

As the use of a low specific tool to identify antibodies against SARS CoV-2 could lead to overestimate the prevalence of SARS CoV-2 infection in settings in which the burden of illness is low or unknown^16^, a more accurate evaluation of which LFAs offer the highest reliability in identifying previously infected individuals or in monitoring the effective response of the immune system to vaccination, could be helpful to select an effective and cheap tool to be used in challenging diagnostic settings.

In this regard, our laboratory evaluated the performance of 11 different LFAs and one ELISA in detecting SARS-CoV-2 infection, analysing plasma and serum samples collected from 350 subjects.

Moreover, 57 plasma samples were collected at 21-24 days from the first dose of BNT162b2 Pfizer-BioNTech vaccine and were examined using the 11 different LFAs and an Electro-ChemiLuminescence ImmunoAssay (ECLIA) dosing IgG against Spike protein Receptor Binding Domain (RBD).

## Material and methods

### Study design: setting and population

This study comprehends two different sampling phases that took place respectively between April - June 2020 and January-February 2021 at San Raffaele Research Hospital in Milan, Italy.

During the first phase, in the spring of 2020, 128 symptomatic COVID-19 patients who resulted positive to rRT-PCR performed on NasoPharyngeal Swab (NPS), participated to the study and from 45 of them two samples were collected at different time points. All patients were hospitalized. Their symptoms included cough (58.13%), dyspnoea (55.40%), and fever (89.01%). Moreover, 49.71% of the patients developed Acute Respiratory Distress Syndrome (ARDS) and 16.18% died. None of the COVID-19 patients had a history of Chronic Obstructive Pulmonary Disease (COPD), but other chronic diseases such as diabetes and hypertension were reported (respectively 9.82% and 32.94%). All clinical data were extracted from the San Raffaele Research Hospital internal database. At the same time point, 26 plasma samples were collected from volunteers who tested negative for SARS-CoV-2 infection by rRT-PCR performed on NPS (denominated in tables and figures as PostH group).

Moreover, to better evaluate the specificity of the tests, 196 samples collected and stored before 2019 were included into the analysis: 82 were from patients in therapy for tuberculosis (TB) and 114 from healthy donors (in tables and figures they are referred to as respectively PreK and PreH group).

All samples were collected by venepuncture, stored at + 4°C and aliquoted for freezing at −80 °C within 1 week of the blood draw. Serum and plasma were used interchangeably for all tests, except for Euroimmun, ELISA, applicable only on serum samples.

The clinical and demographic characteristics of this population are summarised in Table 1 and 2.

**Table 1.** Demographics and clinical characteristics of COVID-19 positive population.

| Variables |  | C-19 POS<br>≤7 days<br>(n=45) |  | C-19 POS<br>8-14 days<br>(n=55) |  | C-19 POS<br>15-35<br>(n=26) |  | C-19 POS<br>Day 36+<br>(n=47) |  |
| --- | --- | --- | --- | --- | --- | --- | --- | --- | --- |
|  |  | N | % | n | % | n | % | N | % |
| Sex | Male | 28 | 62.22 | 38 | 69.09 | 19 | 73.08 | 33 | 70.21 |
|  | Female | 17 | 37.78 | 17 | 30.91 | 7 | 26.92 | 14 | 29.79 |
|  | NA | 0 | NA | 0 | NA | 0 | NA | 0 | NA |
| Ethnic Group | Caucasian | 36 | 80 | 45 | 81.82 | 22 | 84.62 | 38 | 80.85 |
|  | Hispanic | 5 | 11.11 | 8 | 14.55 | 4 | 15.38 | 6 | 12.77 |
|  | Asian | 2 | 4.44 | 1 | 1.82 | 0 | 0 | 1 | 2.13 |
|  | Black | 2 | 4.44 | 1 | 1.82 | 0 | 0 | 2 | 4.26 |
|  | NA | 0 | NA | 0 | NA | 0 | NA | 0 | NA |
| Hypertension | no | 29 | 64.44 | 35 | 63.64 | 17 | 65.38 | 35 | 74.47 |
|  | yes | 16 | 35.56 | 20 | 36.36 | 9 | 34.62 | 12 | 25.53 |
|  | NA | 0 | NA | 0 | NA | 0 | NA | 0 | NA |
| Coronary Artery Disease | no | 42 | 93.33 | 48 | 87.27 | 25 | 96.15 | 46 | 97.87 |
|  | yes | 3 | 6.67 | 7 | 12.73 | 1 | 3.85 | 1 | 2.13 |
|  | NA | 0 | NA | 0 | NA | 0 | NA | 0 | NA |
| Diabetes | no | 35 | 77.78 | 44 | 80 | 20 | 76.92 | 43 | 91.49 |
|  | yes | 4 | 8.89 | 5 | 9.09 | 5 | 19.23 | 3 | 6.38 |
|  | NA | 6 | 13.33 | 6 | 10.91 | 1 | 3.85 | 1 | 2.13 |
| COPD | no | 45 | 100 | 55 | 100 | 26 | 100 | 47 | 100 |
|  | yes | 0 | 0 | 0 | 0 | 0 | 0 | 0 | 0 |
|  | NA | 0 | NA | 0 | NA | 0 | NA | 0 | NA |
| Neoplasia | no | 44 | 97.78 | 53 | 96.36 | 26 | 100 | 47 | 100 |
|  | yes | 1 | 2.22 | 2 | 3.64 | 0 | 0 | 0 | 0 |
|  | NA | 0 | NA | 0 | NA | 0 | NA | 0 | NA |
| Cough | no | 22 | 48.89 | 22 | 40.00 | 10 | 38.46 | 18 | 38.3 |
|  | yes | 23 | 51.11 | 32 | 58.18 | 16 | 61.54 | 29 | 61.7 |
|  | NA | 0 | 0 | 1 | 1.82 | 0 | 0 | 0 | 0 |
| Dyspnoea | no | 19 | 42.22 | 21 | 38.18 | 13 | 50 | 24 | 51.06 |
|  | yes | 26 | 57.78 | 34 | 61.82 | 13 | 50 | 23 | 48.94 |
|  | NA | 0 | NA | 0 | NA | 0 | NA | 0 | NA |
| Fever | no | 7 | 15.56 | 5 | 9.09 | 3 | 11.54 | 4 | 8.51 |
|  | yes | 38 | 84.44 | 50 | 90.91 | 23 | 88.46 | 43 | 91.49 |
|  | NA | 0 | NA | 0 | NA | 0 | NA | 0 | NA |
| Gastro-Intestinal | no | 41 | 91.11 | 47 | 85.45 | 24 | 92.31 | 44 | 93.62 |
|  | yes | 4 | 8.89 | 7 | 12.73 | 2 | 7.69 | 3 | 6.38 |
|  | NA | 0 | 0 | 1 | 1.82 | 0 | 0 | 0 | 0 |
| Headache | no | 44 | 97.78 | 51 | 92.73 | 25 | 96.15 | 44 | 93.62 |
|  | yes | 1 | 2.22 | 3 | 5.45 | 1 | 3.85 | 3 | 6.38 |
|  | NA | 0 | 0 | 1 | 1.82 | 0 | 0 | 0 | 0 |
| Syncope | no | 42 | 93.33 | 52 | 94.55 | 26 | 100 | 46 | 97.87 |
|  | yes | 3 | 6.67 | 2 | 3.64 | 0 | 0 | 1 | 2.13 |
|  | NA | 0 | 0 | 1 | 1.82 | 0 | 0 | 0 | 0 |
| Ageusia-Anosmia | no | 43 | 95.56 | 53 | 96.36 | 26 | 100 | 46 | 97.87 |
|  | yes | 2 | 4.44 | 1 | 1.82 | 0 | 0 | 1 | 2.13 |
|  | NA | 0 | 0 | 1 | 1.82 | 0 | 0 | 0 | 0 |
| Myalgia-Arthralgia | no | 44 | 97.78 | 50 | 90.91 | 26 | 100 | 45 | 95.74 |
|  | yes | 1 | 2.22 | 4 | 7.27 | 0 | 0 | 2 | 4.26 |
|  | NA | 0 | 0 | 1 | 1.82 | 0 | 0 | 0 | 0 |
| Chest Pain | no | 38 | 84.44 | 53 | 96.36 | 26 | 100 | 45 | 95.74 |
|  | yes | 7 | 15.56 | 1 | 1.82 | 0 | 0 | 2 | 4.26 |
|  | NA | 0 | 0 | 1 | 1.82 | 0 | 0 | 0 | 0 |
| ARDS | no | 19 | 42.22 | 21 | 38.18 | 10 | 38.46 | 21 | 44.68 |
|  | yes | 23 | 51.11 | 30 | 54.55 | 10 | 38.46 | 23 | 48.94 |
|  | NA | 3 | 6.67 | 4 | 7.27 | 6 | 23.08 | 3 | 6.38 |
| Death | no | 34 | 75.56 | 42 | 76.36 | 25 | 96.15 | 44 | 93.62 |
|  | yes | 11 | 24.44 | 13 | 23.64 | 1 | 3.85 | 3 | 6.38 |
|  | NA | 0 | NA | 0 | NA | 0 | NA | 0 | NA |
| ICU | no | 31 | 68.89 | 34 | 61.82 | 15 | 57.69 | 37 | 78.72 |
|  | yes | 14 | 31.11 | 21 | 38.18 | 11 | 42.31 | 10 | 21.28 |
|  | NA | 0 | NA | 0 | NA | 0 | NA | 0 | NA |

**Table 2.**
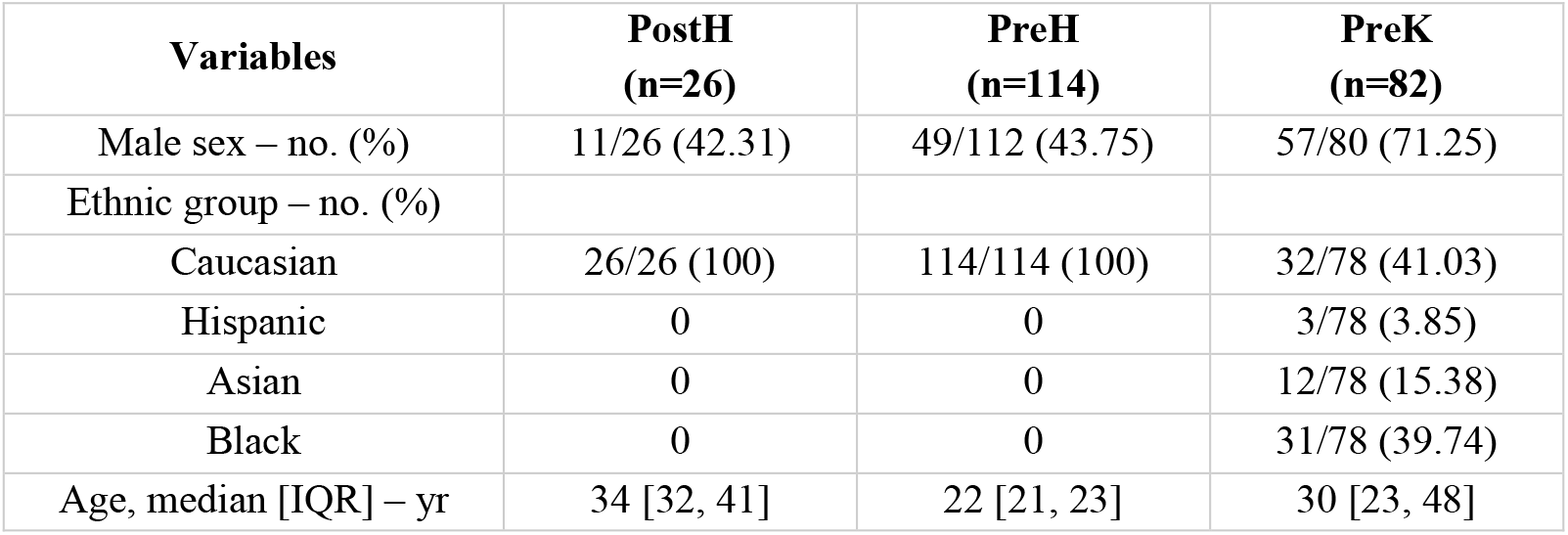
Demographics of COVID-19 negative population.

| Variables | PostH<br>(n=26) | PreH<br>(n=114) | PreK<br>(n=82) |
| --- | --- | --- | --- |
| Male sex – no. (%) | 11/26 (42.31) | 49/112 (43.75) | 57/80 (71.25) |
| Ethnic group – no. (%) |  |  |  |
| Caucasian | 26/26 (100) | 114/114 (100) | 32/78 (41.03) |
| Hispanic | 0 | 0 | 3/78 (3.85) |
| Asian | 0 | 0 | 12/78 (15.38) |
| Black | 0 | 0 | 31/78 (39.74) |
| Age, median [IQR] – yr | 34 [32, 41] | 22 [21, 23] | 30 [23, 48] |

Moreover, a cohort of 57 health workers who had received the first dose of BNT162b2 Pfizer-BioNTech vaccine 21 to 24 days before, was surveyed between January and February 2021. They were all adults, females and males were equally represented. None of them reported any relevant comorbidity or previous immunological disease or allergic reaction to drugs and/or vaccines. From this cohort, all samples were collected by venepuncture, stored at + 4°C and analysed in the 24 hours following the collection using 11 different LFAs and an Electro-ChemiLuminescence ImmunoAssay (ECLIA) dosing IgG against Spike protein RBD.

### Immunochromatographic LFAs

Eleven LFAs were utilized, according to manufacturer instructions (Supplementary Table 1).

In brief, the appropriate sample volume was added on the indicated sample port, followed by a defined amount of the provided diluent. The cartridges were then incubated at room temperature for the recommended time. The results’ reading was performed by two independent observers. Samples were considered negative if the control band was present and the test band absent, positive if both the bands were clearly observed, indeterminate if the control band was present jointly to a faint test band and invalid if the control band was not identified.

### ELISA

Euroimmun Anti-SARS-CoV-2 ELISA for the detection of IgG against SARS-CoV-2 S1 domain was carried out according to manufacturer instructions. In brief, 10 μl of serum were diluted 1:101 in the provided sample buffer. Then 100 μl of the diluted samples, calibrator and positive and negative controls were transferred into the precoated microplate wells according to the provided pipetting protocol and incubated at 37°C for 60 minutes. Following the washing step, conjugate and then substrate incubations were performed before the addition of the stopping solution and the consequent photometric measurement.

### ECLIA

Elecsys® Anti-SARS-CoV-2 S (Roche) is an Electro-chemiluminescence immunoassay (ECLIA) immunoassay for the determination of IgG against SARS-CoV-2 spike (S) protein receptor binding domain (RBD). The assay, based on a double-antigen sandwich assay format has been performed according to manufacturer instruction on Cobas e 411 analyzer on the 57 samples collected from health workers after the first vaccination dose.

### Statistical analysis

Descriptive statistics of continuous variables were presented as median and interquartile range, while for categorical variables frequencies were reported. In the absence of a gold-standard test for serology detection, sensitivity and specificity were estimated using surrogate reference standards. Sensitivity was estimated using samples collected from patients confirmed by rRT-PCR to have been infected with SARS-CoV-2, while specificity was estimated using samples from healthy negative controls and patients receiving therapy for tuberculosis collected prior to the circulation of SARS-CoV-2. Binomial exact 95% confidence intervals were calculated for all estimates. Logistic mixed-effects models were used to evaluate differences among groups, since the data consist of repeated measurements of the same subjects. The agreement between assays was evaluated by computing the percentage of concordant results. All statistical analyses were performed using R statistical software (version 4.0.4; www.r-project.org).

### Ethical approvals

This study was approved by the ethical committee and institutional review board of San Raffaele Research Hospital in Milan, Italy. All patients and healthy controls agreed to the study by signing the informed consent.

## Results

### Test performance

Six out of the eleven analysed LFAs demonstrated perfect specificity in healthy negative controls collected before 2019 (PreH) for both IgM and IgG. BTNX, QuickZen and Tigsun identified one PreH sample as IgM positive (0.89%, 95% C.I. 0.02-4.87); Perfectus and Tigsun identified two samples as IgG positive (respectively 1.75 % 95% C.I. 0.21-6.19 and 1.77 %, 95% C.I. 0.22-6.25) and Right Sign one as IgG positive (0.88%, 95% C.I. 0.02-4.83).

The number of false positives increased dramatically in the group of samples collected before 2019 from patients receiving therapy for tuberculosis (PreK). There was at least one indeterminate or false positive for IgM and/or IgG for these samples across all LFAs (Fig. 1).

**Figure 1.**
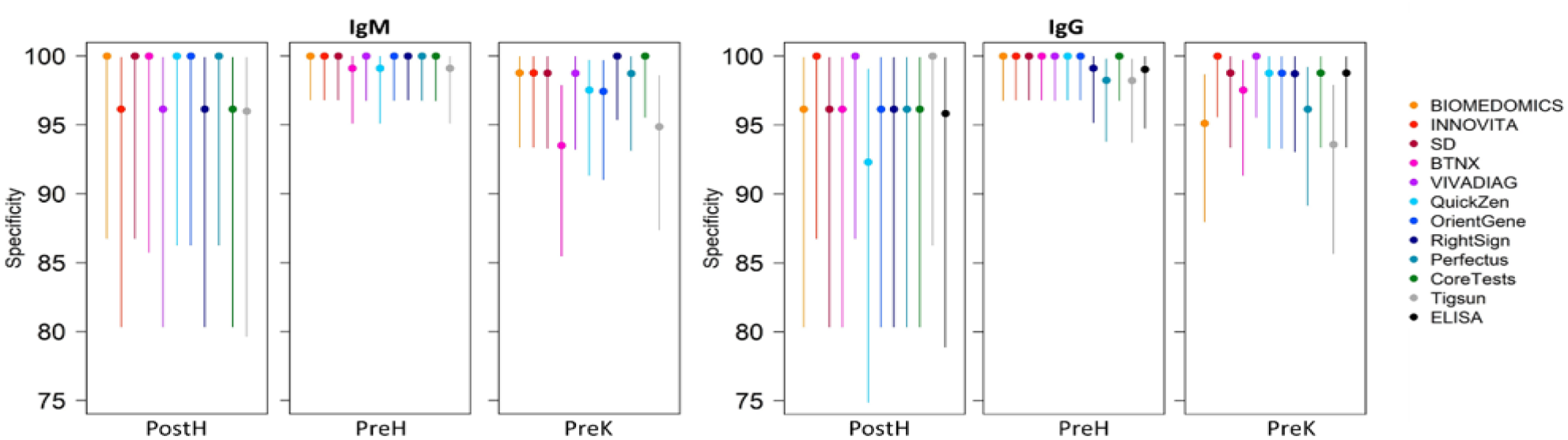
Specificity of ELISA and LFAs calculated evaluating the number of negatives identified in true negative samples groups. PreH: healthy donors sampled before 2019; PreK: patients in therapy for tuberculosis collected before 2019; PostH: volunteers negative to SARS-CoV-2 rRT-PCR performed on nasopharyngeal sample surveyed in 2020

In PostH cohort LFAs’ specificity ranged from 96.15% to 100% for IgM and from 92.31% to 100% for IgG. ELISA demonstrated a specificity of 95.83% (CI 95% 78.88-99.89).

Logistic mixed-effects models were used to evaluate the differences among PreH, PostH and PreK groups. For IgM, the analysis showed that the results were significantly less likely to be negative in TB subjects (PreK) compared to healthy subjects (PreH) (OR=0.12, p-value=0.0115). For IgG, no statistically significant differences have been observed.

In the models, the effect of sex, age and ethnicity was also assessed, and no differences have been observed. The group analysed, including COVID-19 patients and negative controls, had a median age of 33 years old (from 18 to 84 years old); males and females were respectively 59% and 41%; 79.8% were Caucasian, 6.1% Hispanic, 4.3% Asian and 9.8% Black.

As different times of seroconversion have been reported in literature^17^, the sensitivity of the tests was assessed in samples collected across different periods in relation to the occurrence of the symptoms. The results of the evaluation are shown in Figure 2.

**Figure 2.**
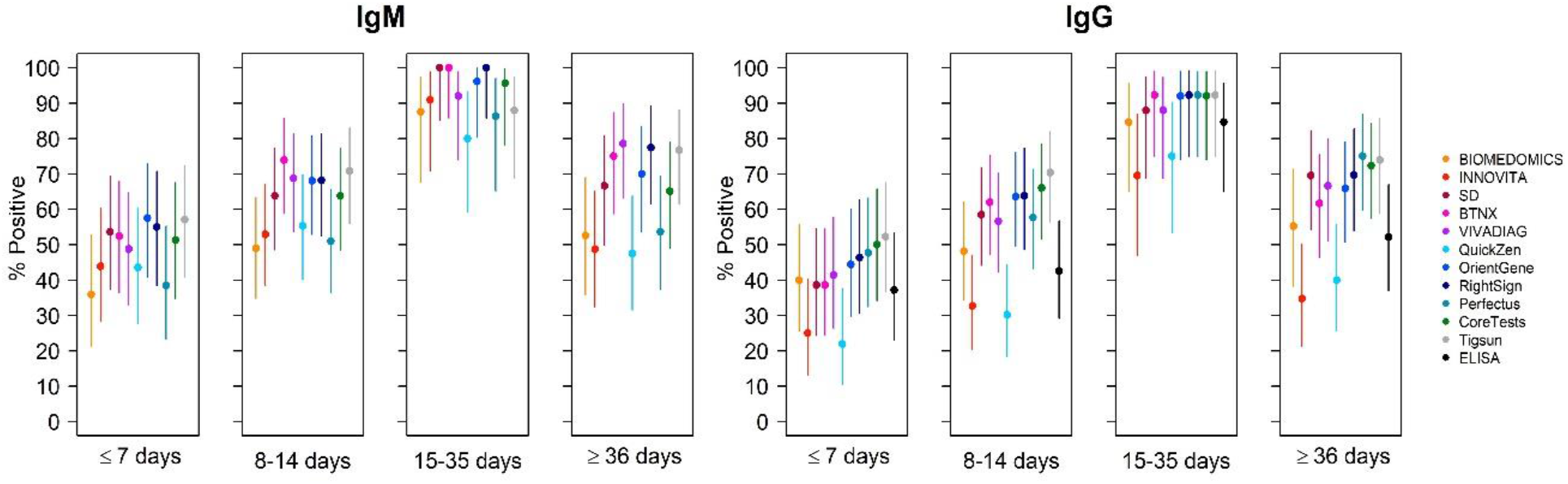
Sensitivity of ELISA and LFAs calculated considering the capability of the test of identifying patients positive for SARS-CoV-2 at different time points after symptoms onset.

Of the six tests that demonstrated perfect specificity in PreH group (Biomedomics, Innovita, SD, VivaDiag, OrientGene, and CoreTests), OrientGene demonstrated the highest sensitivity for IgM at <7 days from symptoms onset (57.50%; 95% CI 40.89-72.96) and CoreTests for IgG (50%; 95% CI 34.19-65.81). Between 8 and 14 days from symptoms onset, VivaDiag had the highest IgM sensitivity of 68.75% (95% CI 53.75-81.34) and CoreTests the highest IgG sensitivity of 66.04% (95% CI 51.73-78.48). At 15-35 days SD had a sensitivity of 100% for IgM (95% CI 85.18-100) and CoreTests of 92.31% for IgG (95% CI 73.97-99.02). Finally, VivaDiag had still a 78.57% IgM sensitivity at more than 36 days from symptoms onset (95% CI 63.19-89.70) and CoreTests identified 72.34% (95% CI 57.36-84.38) of the samples in this group as IgG positive.

Overall, LFAs’ capability of identifying individuals with a SARS-CoV-2 infection proven by rRT-PCR performed on NPS who had seroconvert, raised after 7 days from symptoms’ onset. (Figure 2).

### Indeterminate analysis

Indeterminate results were not observed with QuickZen, OrientGene, RightSign, Perfectus, CoreTests and Tigsun. The two tests with the highest number of IgM indeterminate results were BTNX (21/129) and Viavadiag for IgG (8/129). All the indeterminate results were repeated once. Out of the 8 IgG VivaDiag indeterminate once repeated 4 resulted clearly positive, 1 negative and 3 remained indeterminate; out of 21 IgM BTNX indeterminate 2 resulted positive, 1 negative and 18 remained indeterminate for both observers who perform the reading (Fig.3). To evaluate the effect on sensitivity and specificity we evaluate the effect of considering all the indeterminate results respectively as positive or negative. Comparing the obtained results to the sensitivity calculated excluding the indeterminate results, if considering as positive the indeterminate results an increase in sensitivity from 2 to 8% was reported according to the test, and a loss in sensitivity of 2% to 7% if considering them as negative (Fig.4). Variations in specificity (Fig.5) have also been reported, a loss in specificity from 2% to 4% considering indeterminates as positive and an increase of 2% to 3% considering them as negative.

**Fig. 3.**
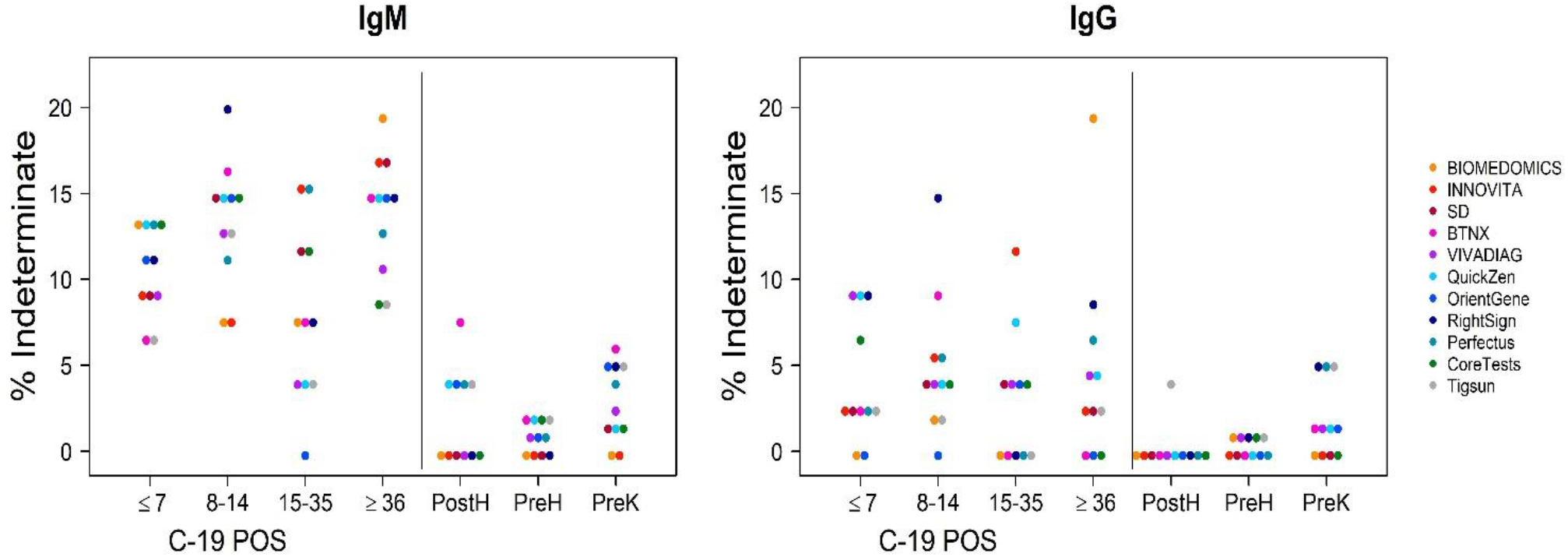
Indeterminates identified by LFAs in the different groups in analysis.

**Fig. 4.**
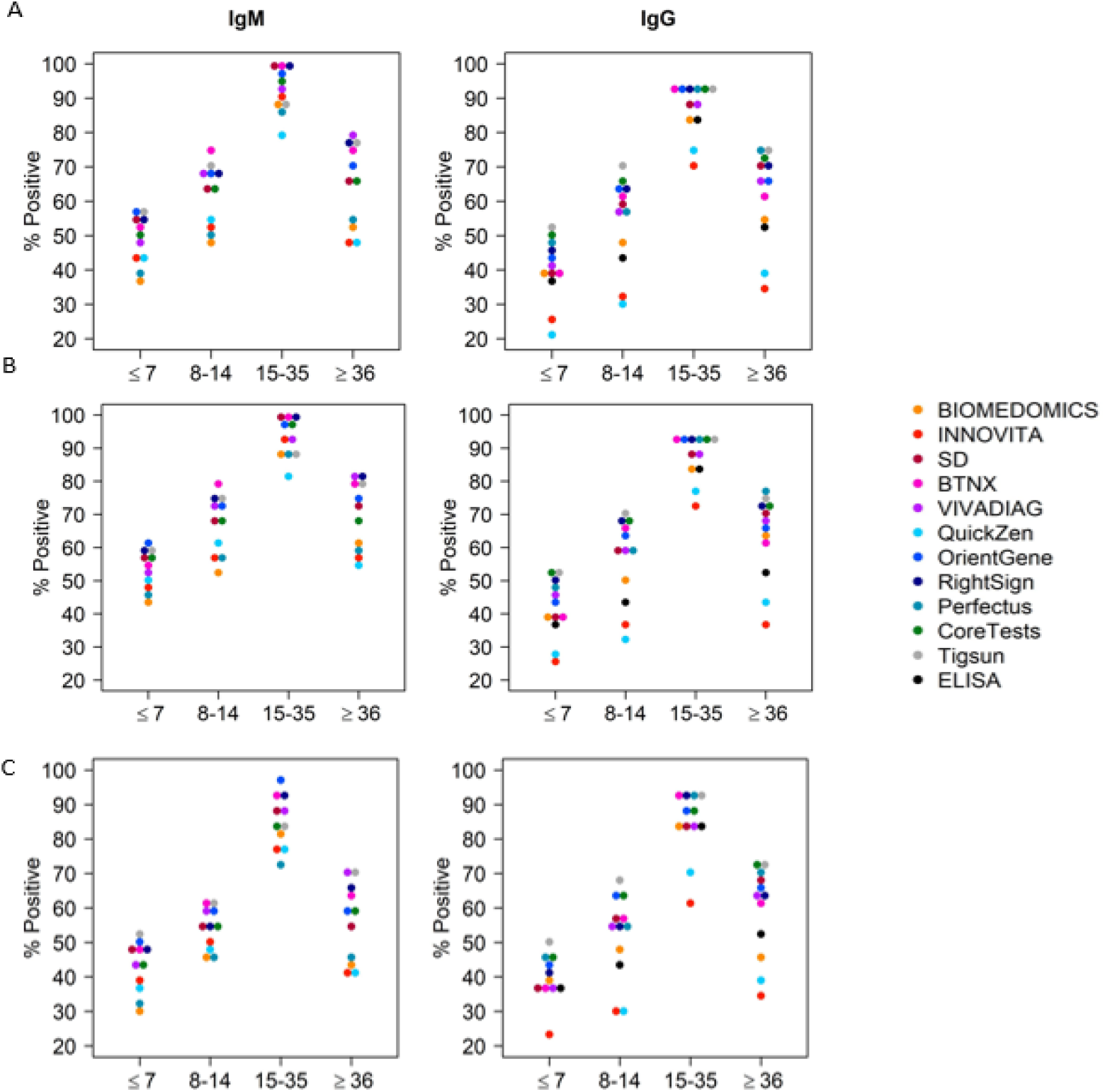
Sensitivity variations considering indeterminate results as positive (b) or negative (c) A: Tests’ sensitivity excluding indeterminate results. B: Sensitivity considering indeterminate results as positive. C: Sensitivity considering indeterminate results as negative.

**Fig. 5.**
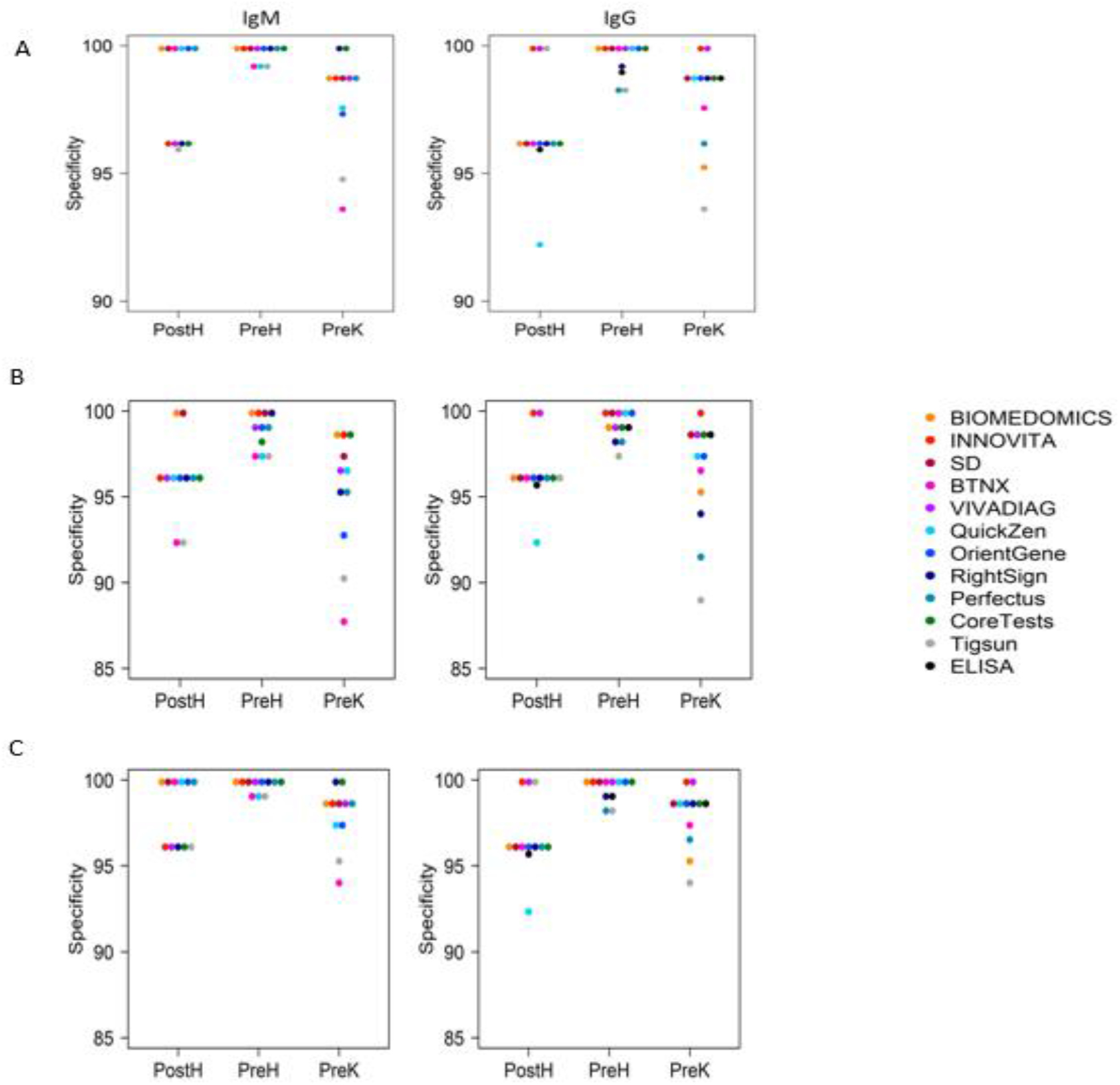
Specificity variations considering indeterminate results as positive (b) or negative (c) A: Tests’ specificity excluding indeterminate results. B: Specificity considering indeterminate results as positive. C: Specificity considering indeterminate results as negative.

### Concordance between tests

The agreement between the different analysed LFAs for IgG and IgM detection and between LFAs and ELISA for IgG has been estimated. In both evaluations the highest concordance rate was observed between 15 and 28 days from the symptoms onset. The concordance level between LFAs and ELISA in the detection of IgG remained at each time point higher than 70%, reaching 100% at 15-35 days for several tests (Fig. 6). The concordance rate between different LFAs IgM and IgG detection in PreH, PostH and PreK groups is provided in Supplementary Fig. 2.

**Fig. 6.**
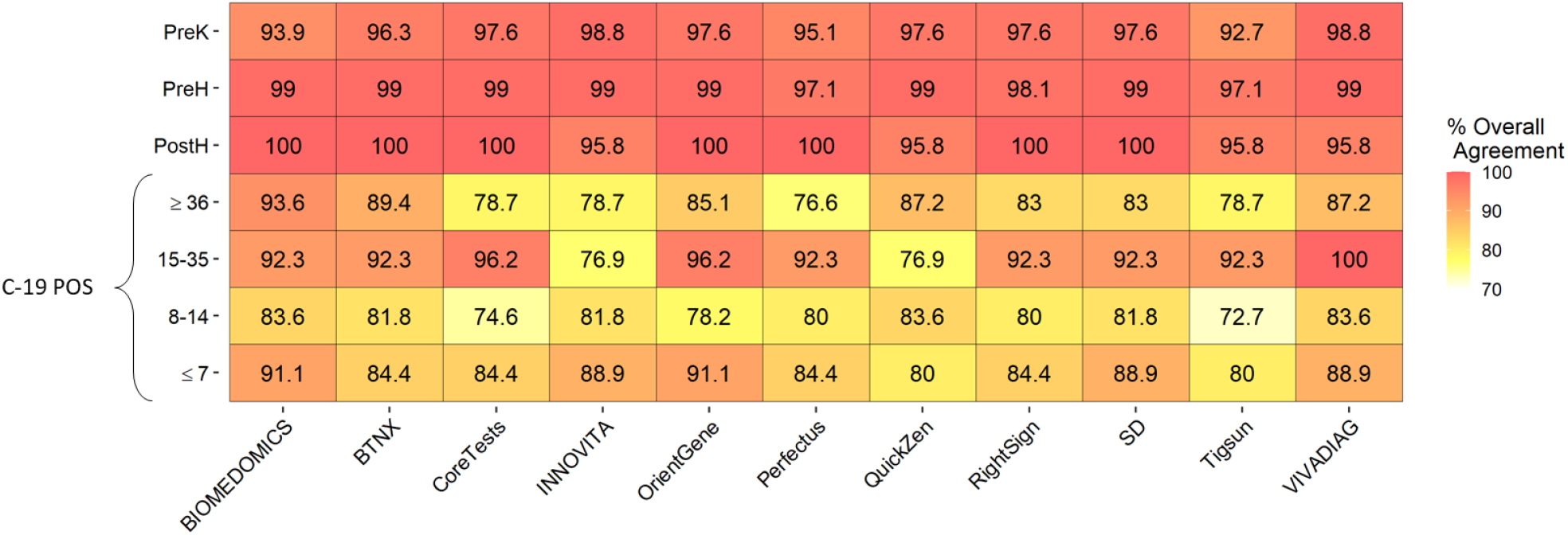
Concordance rate between ELISA and LFAs in different groups and at different time points.

### Seroconversion pattern

An evaluation of the seroconversion pattern has been performed sampling 45 individuals at two different time points. Of 16 patients sampled at ≤ 7 days from symptoms’ onset, 3 were reanalysed between 15 and 35 days and 13 at ≥ 36 days; of the 23 patients firstly sampled between 8 and14 days 3 were again collected between 15 and 35 days and 20 at ≥ 36 days and finally of 6 sampled at 15-35 days and then at ≥ 36 days. Even if a few seroconversions have been observed with all tests in analysis, IgM seroreversion was observed within 30 days from symptoms’ onset with Biomedomics, Innovita, SD, and BTNX (Fig. 7A). Moreover, SD, BTNX, QuickZen, OrientGene, and Perfectus identified each a different sample that reverted for IgG within one month. These results were not confirmed by ELISA, as at this analysis the samples in which the seroreversion was observed resulted or negative or positive at both time points. No seroreversion for IgG was observed at ELISA (Fig. 7B).

**Fig. 7.**
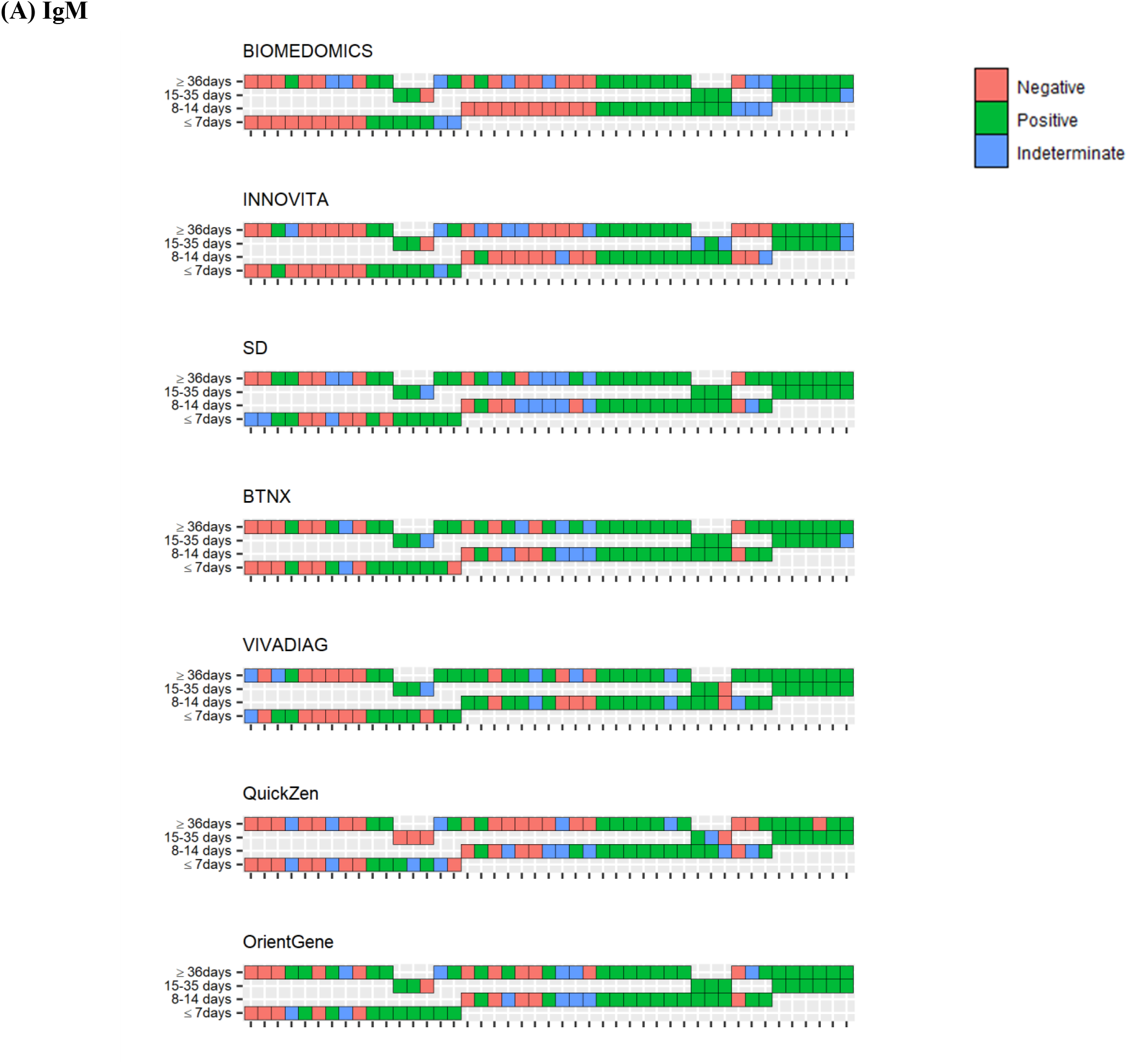
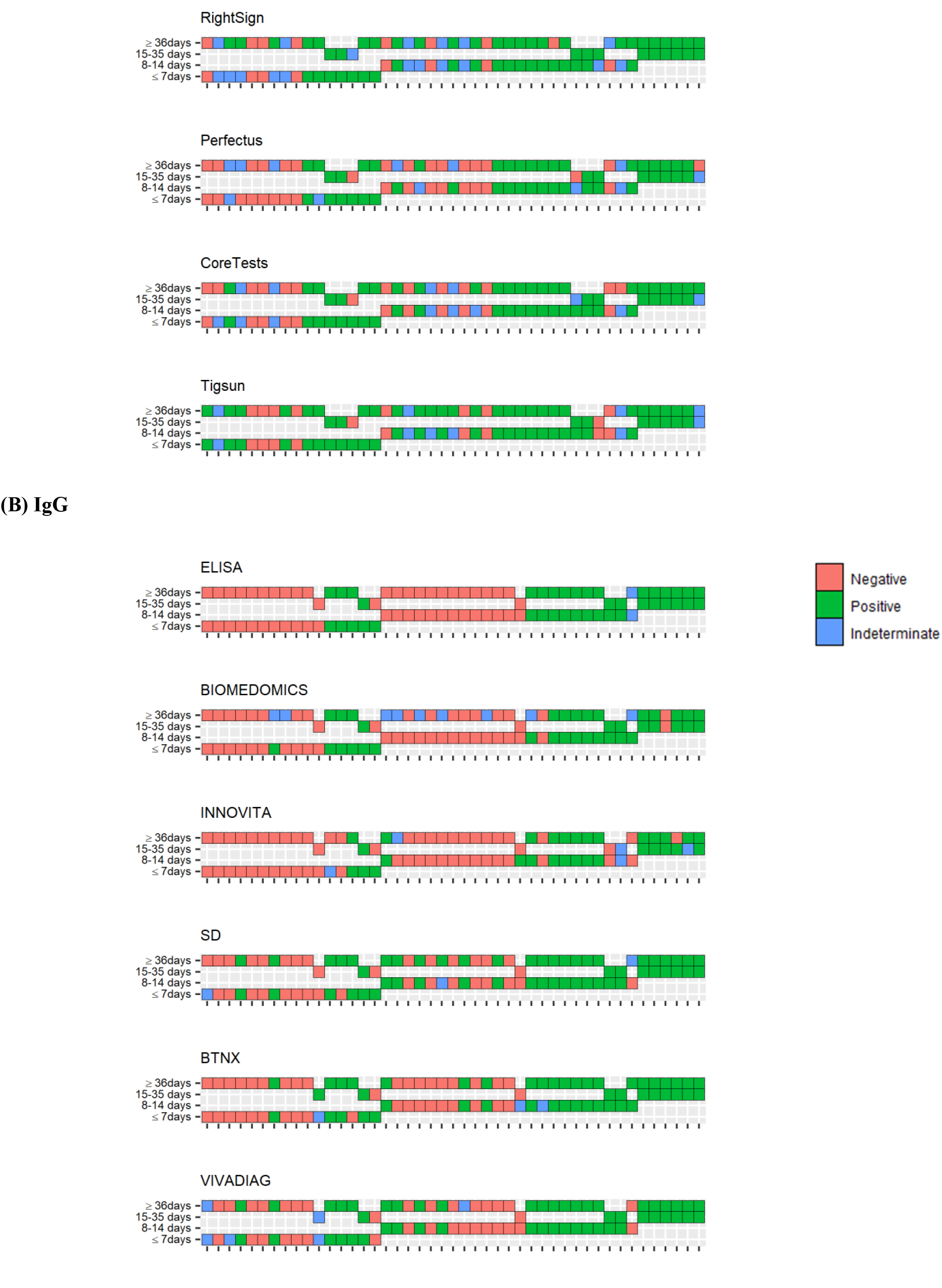
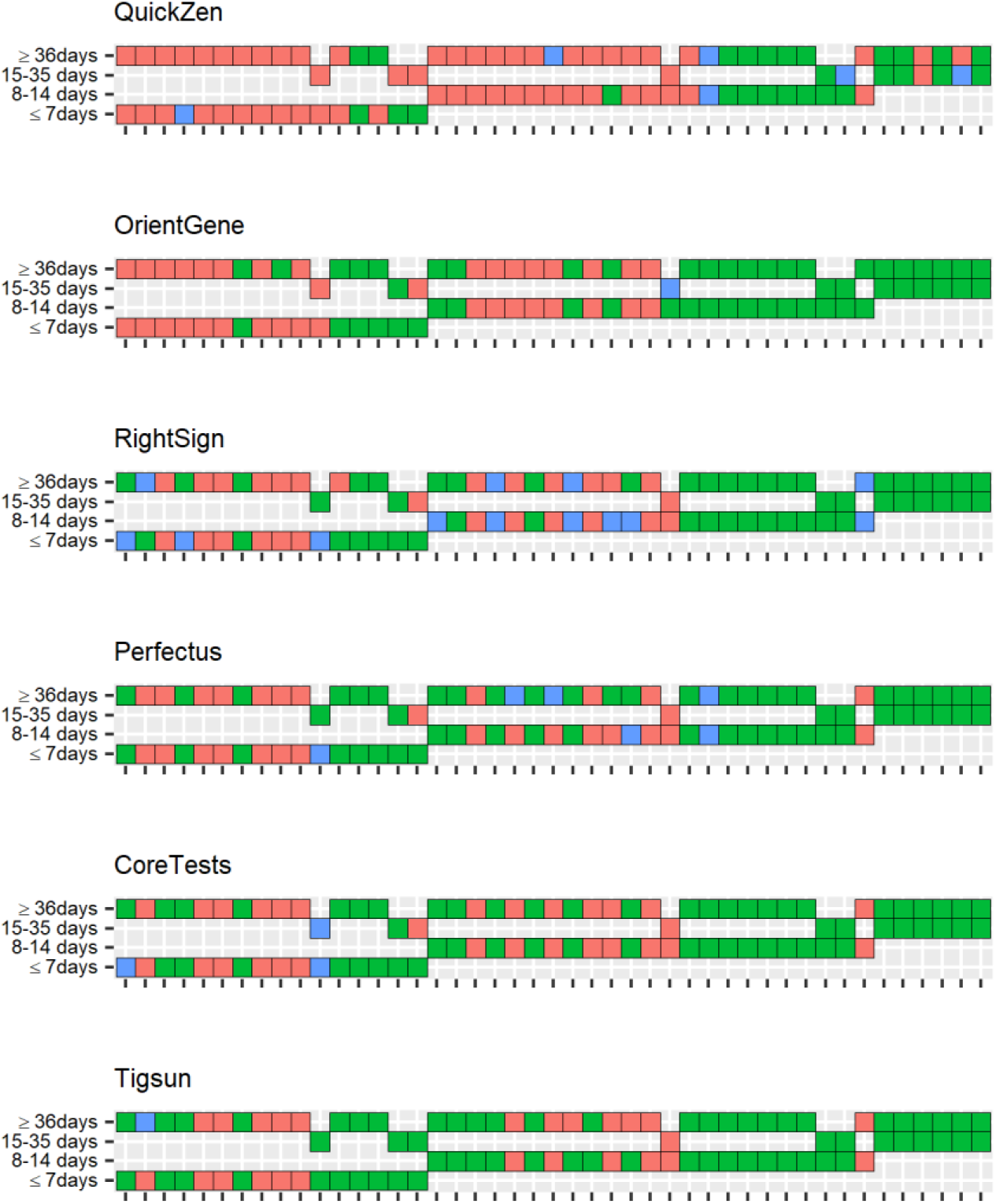
IgM and IgG evaluated at two different time points in the same 45 individuals.

### Healthcare workers

Of the 57 health workers sampled at 21-24 days from the first dose of the Pfizer vaccine, all showed a positive titre of IgG against SARS-CoV-2 Spike protein RBD (≥0.8 U/mL), detectable by ECLIA (Table 3). Of the 11 LFAs used to detect an IgM response, only two, Tigsun and BTNX, failed to show a positive response in any of the samples tested, while OrientGene had a IgM positivity rate of 12.24% (6/49) and the highest IgG detection rate (85.71%) (48/56) (Table 4).

**Table 3.** Quantitative IgG ECLIA results compared with IgG positive or negative LFAs’ identification in vaccinated healthworkers.

| Ab-COVID-Spike (ECLIA) | IgG Negative to LFA |  |  | IgG Positive to LFA |  |  |
| --- | --- | --- | --- | --- | --- | --- |
|  | median | min | Max | median | min | Max |
| VivaDiag | 54.1 | 1.57 | >2500 |  |  |  |
| Innovita | 25.85 | 1.57 | >2500 |  |  |  |
| BTNX | 5.49 | 1.57 | 11.5 | 66.95 | 15.4 | 229 |
| Biomedomics | 12.655 | 6.21 | 19.1 | 62 | 31.8 | 132 |
| SD | 16.5 | 1.57 | 107 | 122 | 5.27 | >2500 |
| CoreTests | 20.45 | 1.57 | 213 | 197 | 5.27 | >2500 |
| RightSign | 17.2 | 1.57 | 107 | 181 | 8.15 | >2500 |
| Perfectus | 11.1 | 1.57 | 107 | 122 | 8.15 | >2500 |
| QuickZen | 19 | 1.57 | 222 | 193 | 11.5 | >2500 |
| OrientGene | 9.74 | 1.83 | 107 | 54.1 | 4.04 | >2500 |
| Tigsun | 36.15 | 1.57 | 367 | 2500 | 5.27 | >2500 |

**Table 4.** Rate of IgM and IgG that resulted positive, negative or indeterminate at different LFAs in vaccinated healthworkers.

| IgM | Negative |  | Positive |  | Indeterminate | Invalid | NA |
| --- | --- | --- | --- | --- | --- | --- | --- |
|  | N | % | N | % |  |  |  |
| VivaDiag | 36 | 97.3 | 1 | 2.7 | 0 | 1 | 20 |
| Innovita | 27 | 96.43 | 1 | 3.57 | 2 | 0 | 28 |
| BTNX | 18 | 100 | 0 | 0 | 2 | 0 | 38 |
| Biomedomics | 5 | 83.33 | 1 | 16.67 | 1 | 0 | 51 |
| SD | 54 | 96.43 | 2 | 3.57 | 2 | 0 | 0 |
| CoreTests | 55 | 98.21 | 1 | 1.79 | 1 | 0 | 1 |
| RightSign | 52 | 92.86 | 4 | 7.14 | 1 | 0 | 1 |
| Perfectus | 52 | 94.55 | 3 | 5.45 | 2 | 0 | 1 |
| QuickZen | 44 | 91.67 | 4 | 8.33 | 8 | 0 | 2 |
| OrientGene | 43 | 87.76 | 6 | 12.24 | 8 | 0 | 1 |
| Tigsun | 57 | 100 | 0 | 0 | 0 | 0 | 1 |
| IgG | Negative |  | Positive |  | Indeterminate | Invalid | NA |
|  | N | % | N | % |  |  |  |
| VivaDiag | 37 | 100 | 0 | 0 | 0 | 1 | 20 |
| Innovita | 30 | 100 | 0 | 0 | 0 | 0 | 28 |
| BTNX | 6 | 37.5 | 10 | 62.5 | 4 | 0 | 38 |
| Biomedomics | 2 | 33.33 | 4 | 66.67 | 1 | 0 | 51 |
| SD | 14 | 28 | 36 | 72 | 8 | 0 | 0 |
| CoreTests | 36 | 67.92 | 17 | 32.08 | 4 | 0 | 1 |
| RightSign | 21 | 47.73 | 23 | 52.27 | 13 | 0 | 1 |
| Perfectus | 15 | 28.85 | 37 | 71.15 | 5 | 0 | 1 |
| QuickZen | 28 | 62.22 | 17 | 37.78 | 11 | 0 | 2 |
| OrientGene | 8 | 14.29 | 48 | 85.71 | 1 | 0 | 1 |
| Tigsun | 50 | 90.91 | 5 | 9.09 | 2 | 0 | 1 |

The rate of positivity to IgG of the different LFAs was evaluated in comparison to the quantitative results obtained by ECLIA. As shown in Table 3 VivaDiag and Innovita did not detect positive IgG neither for ECLIA titres > 2500 U/ml. The lowest positive titre was correctly identified by OrientGene (4.04 U/ml).

## Conclusions

In this study we analysed the performance of 11 different LFAs and one commercial ELISA in detecting SARS-CoV-2-specific IgG and IgM. Specificity was assessed in three cohorts: historic samples collected before 2019 in healthy donors and in patients who were on TB treatment; as well as individuals who tested RT-PCR negative for SARS-CoV-2. The overall sensitivity of the tests (positive to IgM and/or IgG) was calculated evaluating the tests capability to correctly identifying individuals confirmed to have been infected with SARS-CoV-2 by rRT-PCR.The obtained data allowed us to identify CoreTest as the test with highest specificity and sensitivity at ≥ 36 days from symptoms onset. Hence, the latter appeared to be more appropriate for a serological mass screening, due to the lower risk of identify false positive because of the high specificity. Furthermore, OrientGene demonstrated the highest sensitivity in identifying a positive titre of IgG against protein S RBD, proving itself a possible test to evaluate an immunological response after the vaccine.An in-depth evaluation on a wider cohort is needed to assess the effective reliability for this purpose of OrientGene in comparison to other LFAs. Interestingly, VivaDiag and Innovita, even if demonstrated good specificity (both 100%, 95 % C.I. respectively 95.55-100 and 95.60-100) and sensitivity (VivaDiag 94.12%, 95% C.I. 71.31-99.85; Innovita 86.67%, 95% C.I. 59.54-98.34) in identifying positive subjects at 15-28 days from symptoms’ occurrence, did not detect the IgG response at 21 days from the vaccination. Indeed, more information by LFAs’ manufacturers on the antigenic targets of their tests would help to perform a more on point evaluation of these tests. A further study, including more timepoints from symptoms’ onset could be useful to evaluate the reliability of LFAs to identify a previous infection of SARS-CoV-2 at 60 and 90 days from the infection. The main limitation in the sensitivity assessment is due to the lack of asymptomatic and pauci-symptomatic patients in our cohort to perform an evaluation of the rate of positivity in association with the severity of the developed disease.

Interestingly, only one test (BTNX) recognised in its product insert rifampicin, ethambutol and isoniazid as possible interfering substances, but declared that sensitivity and specificity of the test were not affected by these drugs presence at therapeutic concentration in blood. Nonetheless, the level of agreement of BTNX with other LFAs for IgM is the lowest in the PreK group, therefore TB therapy could have had an effect on the test despite what is declared by the manufacturer. Even if it is well known that rifampicin can cause false-positive immunoassay results for urine opiates,^18^ to our knowledge, this is the first report that provide proof that TB medicines can affect SARS-CoV-2 LFAs for antibodies detection. This occurrence probably deserves an in-depth analysis to identify the possible mechanisms for cross-reactivity, but it is to be kept in account if LFAs will be used in countries with a high TB prevalence.

A higher number of indeterminate results was overall observed for IgM than IgG. The identification of these faint bands affected the general efficiency and reliability mainly of two of the LFAs in analysis, BTNX and VivaDiag. The repetition of the indeterminate exams did not provide a clear positive or negative result in the majority of cases for BTNX IgM, as 13/21 still remained not interpretable. Previous studies have suggested to consider all faint identified bands as negative to improve the specificity of the test.^10^ However, our analysis demonstrated that considering negative all the samples identified as indeterminate would result in a major decrease in the sensitivity of the tests (up to 7%) compared with a minimal gain in specificity (2 to 3%). Moreover, the definition of “faint” is based on a subjective evaluation of the band intensity that in future could be objectify through the use of automatic readers or, at least, through an attentive training of the readers. ^19^

In conclusion, the tests analysed demonstrated different performances and different levels of reliability in identifying IgM and IgG against SARS-CoV-2. Therefore, great prudence should be used to employ the most accurate POC serological tests to evaluate the local epidemiology as well as for verifying the development of an immunological response after the vaccine, especially in diagnostic challenging settings. The need for readers’ training as well as the possible interference of TB therapy on the tests results have been identified by our study as two of the main limiting factors for the use of these tests in Low-Middle Income Countries. Finally, in a period of scarcity of vaccines’ doses, when several European countries, including Italy and France, are recommending a single dose of vaccine for individuals who were positive for SARS-CoV-2 in the previous six months, the tests with the highest specificity could be used to determine a prior infection and therefore deeply influence the vaccination campaign.^3,4^

## Data Availability

Not external dataset or other repositories.

---

**Supplementary Table 1.** LFAs manufacturers and Antigenic target.

| Name | Manufacturer | Ag Target |
| --- | --- | --- |
| COVID-19 IgM/IgG Rapid Test | Biomedomics | Undisclosed |
| 2019-nCoV Ab Test | Innovita | S1+N |
| STANDARD Q COVID-19 IgM/IgG Duo | SD Biosensor | N |
| COVID-19 IgG/IgM Test | BTNX | Undisclosed |
| COVID19 Single Rapid Test IgM/IgG | VivaDiag | Undisclosed |
| COVID-19 IgM/IgG | QuickZen | Undisclosed |
| COVID-19 IgG/IgM Rapid Test | OrientGene | Undisclosed |
| COVID-19 IgG/IgM Rapid Test Cassette | RightSign | Undisclosed |
| Novel Corona Virus (SARS-CoV-2) IgM/IgG Rapid Test Kit | Bio Perfectus | Undisclosed |
| COVID-19 IgM/IgG Ab Test | CoreTests, Core Technology | Undisclosed |
| COVID-19 Combo IgM/IgG Rapid Test (Lateral Flow Method) | Tigsun | Undisclosed |
| Anti-SARS-CoV-2 ELISA IgG | Euroimmun | S1 |

**Supplementary Figure 2.**
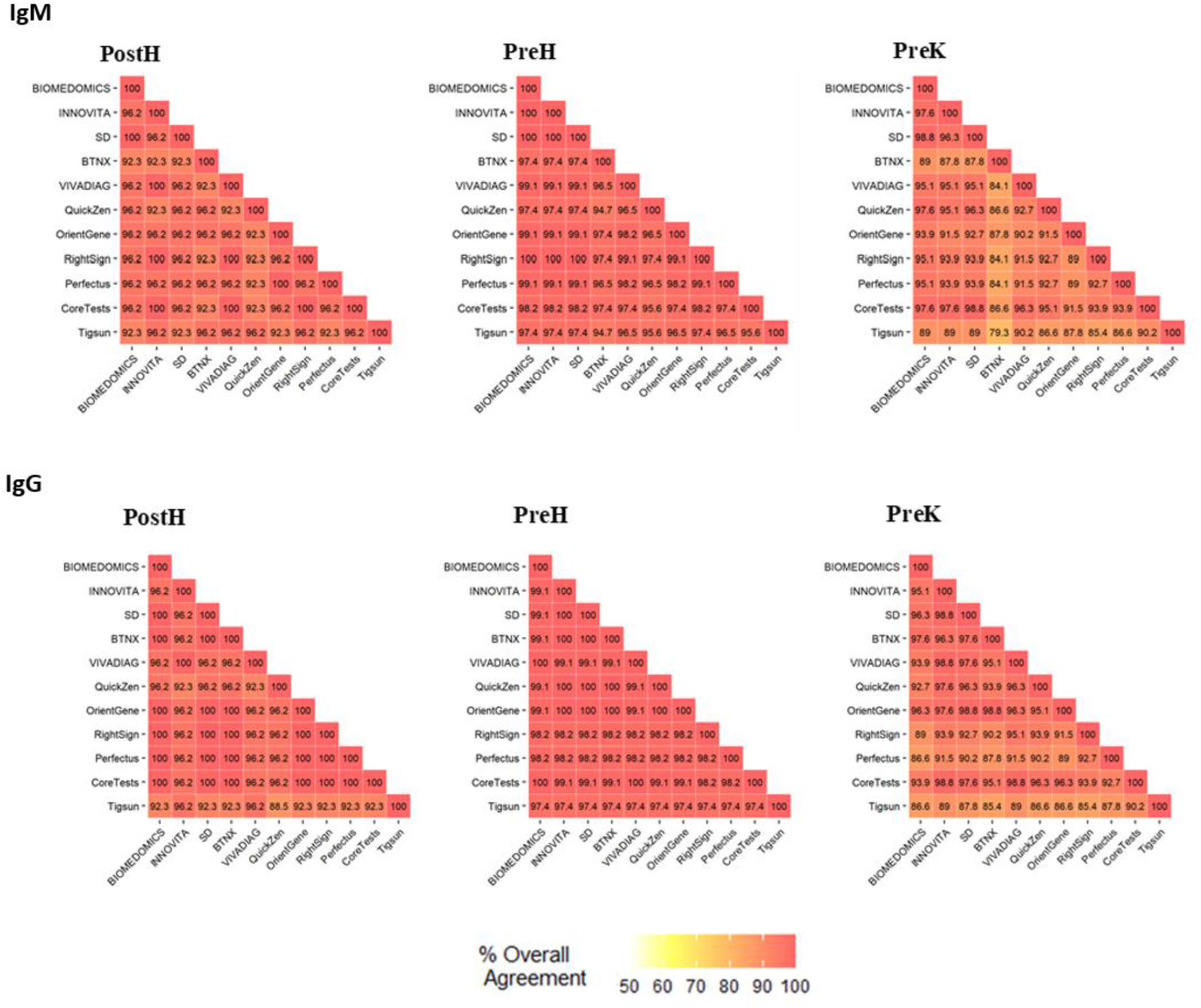
Accordance between LFAs.

## Notes

### Competing Interest Statement

The authors have declared no competing interest.

### Funding Statement

Foundation for Innovative New Diagnostics (Paola Mantegani)
World Health Organization, Unitaid, UK Department for International Development (Margaretha De Vos)

### Author Declarations

San Raffaele Scientific Institute Ethics committee

